# Tactile Perception and Tolerability Thresholds of TMS Characterized by Intensity across Locations and Frequencies

**DOI:** 10.64898/2026.01.28.26345065

**Authors:** Neketa Nesmith, Megan Senda, Yang Hou, Kirthin Dev, Austin M. Spitz, F. Andrew Kozel, Kevin A. Johnson

## Abstract

Transcranial Magnetic Stimulation (TMS) involves pulsed magnetic fields that pass through the scalp to stimulate the brain, with incidental stimulation to superficial nerves and muscles. From a research perspective, the tactile sensations can be a problematic confound, particularly when stimulation approaches an unpleasant or painful level. Additionally, tactile sensations contribute to difficult challenges in establishing an appropriate sham control condition. Clinically, some patients find stimulation uncomfortable or intolerable. Clinicians need data on adjustments to stimulation parameters to improve tolerability and efficacy. The primary objective of this study was to characterize the tolerability of TMS by location (over modified Beam F3 prefrontal, THREE-D prefrontal, right orbitofrontal, medial prefrontal, motor, and parietal cortical targets, as well as the knee) and by frequency (1 Hz, 10 Hz, or iTBS), with increasing levels of stimulation intensity. We also characterized sensory thresholds and qualitative aspects of stimulation across locations and frequencies. For location, sites distal to the facial nerves and muscle (Knee, P3, M1, mPFC) were more tolerable, followed by Beam F3, with the THREE-D and AF8 locations as least tolerable. For frequency, we found that 1 Hz was significantly more tolerable than 10 Hz and iTBS. iTBS was more annoying than 10 Hz but only marginally different in tolerability. TMS researchers and clinicians should understand the impact of sensation based on location and frequency, with increasing stimulation intensity. This is a single-session study in generally healthy individuals, and there is a need for additional data to further inform research and clinical practice.

## INTRODUCTION

Transcranial Magnetic Stimulation (TMS) involves pulsed magnetic fields that pass through the scalp to stimulate the brain, with incidental stimulation to superficial nerves and muscles [1–4]. As the magnetic field decays exponentially from the TMS coil, the energy impacting peripheral nerves is greater than the energy reaching the brain cortex [5, 6]. Researchers and clinicians primarily focus on the brain effects of TMS, which vary based on location and frequency of stimulation [7, 8]. The tactile sensations of TMS also vary based on frequency and location [2, 9], as well as pulse width and the stimulation strength that is typically individually set based on motor threshold (MT) [10]. Individual anatomy impacts the sensory experience related to the variability of stimulation intensity (set by MT) and positioning [11–13]. Psychological factors such as anxiety and appraisal can also impact tactile perceptions of pain [14, 15]. Experienced TMS users know that that tolerability varies by scalp location (e.g., sites closer to facial muscles and cranial nerves are perceived as less tolerable), although a full understanding is limited by relatively sparse data [2, 16]. Experienced users of TMS also informally discuss that stimulation tolerability varies by frequency (e.g., 1 Hz is clearly more tolerable than 10 Hz, and 10 Hz may be more tolerable than intermittent theta-burst, iTBS), with only limited empirical data to support assumptions [9, 14]. Formal quantitative evaluations of multiple frequencies at multiple locations are a gap in the research literature. Given the significant implications for research and clinical applications of TMS, we sought to develop methods and characterize tactile sensations associated with common scalp locations targets and stimulation frequencies.

From a neuroscience methods perspective, the tactile sensations can be a challenging confound, particularly when stimulation approaches an unpleasant or painful level [17]. Researchers often use TMS to probe foundational neuroscience questions including motor physiology and cognitive processing [18, 19]. Painful peripheral sensations can alter corticomotor excitability and MT [20, 21] and can also impact attention and cognitive processing [2, 22, 23]. Tactile sensations may become a confound when the pain or discomfort differs between comparisons, including different frequencies, scalp locations, or individual intensities set by MT. Physiological measures, such as heart rate deceleration in frontal locations versus acceleration in non-frontal control locations, have been proposed as biomarkers of brain target engagement [24, 25]. Noxious sensory stimuli also impact cardiac autonomic responses [26, 27], potentially confounding interpretation of brain versus tactile effects, especially with perceptual differences by location and intensity.

From a clinical research perspective, the tactile sensations of TMS contribute to the difficult challenge in establishing the appropriate sham control condition [28–30]. A sham without tactile sensation risks unblinding participants, while a sham with tactile sensation may have physiological effects that contribute to therapeutic benefit.

Neuromodulation of cranial nerves has been explored and indicated for treating neurologic and psychiatric disorders [31, 32]. Additionally, noxious stimulation can activate endogenous opioid systems that can impact mood [33–35]. Controls comparing different frequencies or locations can be challenging to interpret if there are tactile differences confounding the comparisons. The THREE-D study is a notable study that established theta-burst stimulation as equivalent to 10 Hz stimulation for treating depression [36]. The study used a different mean location from the OPT-TMS study [11, 37], and there was a high rate of reported headache in both THREE-D groups. TMS clinical studies have not provided standardized, detailed information about tactile discomfort (e.g., intensity, duration) to fully understand the potential impacts.

From a clinical perspective, scalp discomfort is a known side effect [4, 38, 39]. There have been attempts to block the superficial effects, although this has not been translated to routine practice [4, 40]. Fortunately, most patients acclimate to the discomfort and tolerate a full course of treatment [14, 38, 39, 41, 42]. The mechanism of this acclimation could be peripheral physiology, psychological reappraisal and reduced attention, or even affective brain effects [41]. Clinicians may utilize a variety of methods to reduce discomfort, from ramping up machine output intensity gradually, adjusting location and frequency, or using distraction. Considering both the variability in clinical experience and the potential efficacy impact of adjusting treatment parameters for tolerability, there is a need for empirical knowledge to guide clinical decision making.

The primary aim of the present study was to systematically characterize the tolerability of TMS across clinically relevant scalp locations (over modified Beam F3 prefrontal, THREE-D prefrontal, right orbitofrontal, medial prefrontal, motor, and parietal cortical targets) and commonly used stimulation frequencies (1 Hz, 10 Hz, and iTBS), evaluated at increasing levels of stimulation intensity (machine output). A secondary objective was to evaluate the knee as a surrogate location to safely trial the tolerability of TMS parameters. Based on cranial anatomy and prior experience, we hypothesized that TMS will be less tolerable at the THREE-D prefrontal and orbitofrontal locations relative to the modified Beam F3 prefrontal location. Based on experience and limited data, we hypothesized that TMS theta burst will be less tolerable than 10 Hz.

## METHODS

### Participants and Procedures

This study was registered on ClinicalTrials.gov (NCT06354686). We recruited adult volunteers, ages 18 years and over, from the local community. Participants first provided informed consent, as approved by the institutional review board. We then administered a TMS safety screen [43], collected relevant self-report questionnaires and conducted a brief medical history. Exclusion criteria covered contraindications for TMS and conditions that would interfere with sensory testing. We registered the study with the NIH protocol registry and results system (NCT06354686).

Prior to sensory testing, we determined each participant’s motor threshold (MT) using the maximum-likelihood strategy [44, 45] and visual observation of the right hand digits. We never exceeded the individual’s 120% MT in subsequent testing for safety considerations [4, 46]. We used a MagPro R30 TMS System (MagVenture A/S, Farum, Denmark) with a Cool-B70 figure-8 coil. On this system, the population average MT is 39% of the maximum machine output (internal data and manufacturer correspondence). For comparisons across different machines, we define this 39% level as 100% of the population average MT on the system (sMT). So, a setting of 20 (19.5 rounded up) on the study TMS system would equate to 50% sMT (i.e. 50% of the population average MT).

After MT determination, we marked scalp locations on a fabric cap with neuronavigation registration. We then conducted sensory testing with each participant at 7 locations, with 3 frequencies per location (21 trials per participant). We used a hierarchical randomization design, considering order effects, possible local induced sensitivity, and efficiency. We randomly assigned from two different location sequences, and with testing at each selected location, we randomized the three stimulation frequencies. Testing typically occurred over a single session lasting approximately 3 hours.

### Location

We performed sensory testing first on the knee, before testing locations on the scalp. We first tested on the knee to provide the participants with a chance to acclimate to the sensations and rating procedures, as well as to evaluate the knee as a surrogate location for safe sensory testing (no seizure risk). We placed the coil just above the patella, with the handle down parallel with the lower leg.

For the scalp locations, we first registered the participant’s head to the MNI template using Brainsight stereotaxic neuronagivation equipment (Rogue Research Inc., Montreal, Quebec, Canada), based on a combination of anatomic and EEG landmarks. This allowed us to approximate the THREE-D MNI location (-38, 44, 26) [36], as well as estimate the MNI coordinates of the other locations tested. All participants wore a white fabric cap with all target locations marked for scalp testing.

The six scalp locations tested were the left dorsolateral prefrontal cortex by the modified Beam method (“Beam F3”) [47], the left dorsolateral prefrontal cortex by the THREE-D coordinates (“THREE-D”) [36], the right orbitofrontal cortex using Downer methodology (AF8) [48], the medial prefrontal cortex found by the EEG Fz convention (“Fz”), the left primary motor cortex as localized during the MT determination (“M1”), the left parietal cortex using inverted modified Beam measurements (“P3”). For both Beam F3 and THREE-D locations, the coil was oriented 45° from midline, handle pointing posterior, following standard clinical convention for treating major depressive disorder. For the AF8 location, the coil was oriented as referenced [48]. For the Fz location, the coil was oriented along the midline, handle pointing posterior, similar to clinical orientation for treating obsessive-compulsive disorder. For the M1 location, the coil was oriented parallel to the midline, handle pointing posterior. For the P3 location, the coil orientation was flipped from Beam F3, handle pointing anterior. We created two testing orders for scalp locations, avoiding sequential testing of the two dorsolateral prefrontal sites, and participants were randomly assigned to one of the orders.

### Frequency

We tested trains of three different frequencies at each location in randomized order. The 1 Hz train consisted of stimulation ON for 4 seconds, 1 pulse per second, then stimulation OFF for 11 seconds. The 10 Hz train consisted of stimulation ON for 4 seconds, 10 pulses per second, then stimulation OFF for 11 seconds. The iTBS train consisted of stimulation ON for 2 seconds, 5 triplet pulses per second (each pulse consisted of 3 pulses at 50 Hz), then stimulation off for 13 seconds.

### Threshold Testing

Our testing was derived from Quantitative Sensory Testing (QST) method of limits [49] and from the McGill Pain Questionnaire (MPQ) [50]. The method of limits is a rapid way to assess thresholds (sensory or pain) by increasing the stimulus intensity until a threshold is reached. The MPQ contains qualitative descriptors that can be linked to the intensity of discomfort or pain. We focused on the evaluative domain, that increases through “annoying”, “troublesome”, “miserable”, “intense”, to “unbearable”. From clinical experience, we selected “annoying” and “troublesome/miserable” as appropriate levels to gauge discomfort and tolerability. We avoided “unbearable” given practical concerns of headache with repeated testing and ethical concerns.

In a single continuous trial, we used an increasing method of limits to establish a “sensory threshold”, an “annoyance threshold”, and a “tolerance threshold”. In the single trial, we administered two trains of the assigned frequency at 30% sMT (machine output = 11), then two trains at 40% sMT (machine output = 15), and subsequently continued with two trains at each subsequent 10% sMT increment (machine output = 19, 23, 27, etc.) until the tolerance threshold was reached (or 120% of the individual’s motor threshold as a safety limit). We instructed participants to identify the sensory threshold as the first train where a tactile sensation was perceived (noting that tactile sensation should be differentiated from auditory perception). We instructed participants to identify the annoyance threshold, after the sensory threshold was identified, as the first train where the tactile sensation was perceived as annoying (descriptor based on the MPQ, Group 16). We instructed participants to identify the tolerability threshold, after the annoyance threshold was identified, as the first train where the tactile sensation was perceived as troublesome or miserable (descriptors based on the MPQ, Group 16). At the conclusion of each trial, participants also provided a numeric pain rating (0 – 10 scale, 0 being none and 10 being worst) and a broader characterization of the sensation based on MPQ (adjectives from groups 1, 2, 4, 5, 8, 9, 12, 13, 16, 17, and 20).

### Data Analysis

We first examined the distributional properties of the dependent variables at each level of the within-subject factors. Skewness and kurtosis values were inspected to assess departures from normality. Consistent with recommended guidelines (±1.96); [51], all variables demonstrated acceptable normality, with only a few instances of moderate kurtosis (between 2 and 7). We used multilevel modeling (MLM; i.e., mixed effects model) [52, 53]) to test our primary aims, given that MLM is robust to moderate deviations from normality and can appropriately account for correlated observations within participants.

MLM was used to evaluate the effects of stimulation location (Knee, Beam F3, THREE-D, AF8, mPFC, M1, P3) and stimulation frequency (1 Hz, 10 Hz, iTBS) on sensory, annoyance, and tolerability threshold values. Repeated observations across stimulation conditions were nested within individuals, and random intercepts accounted for individual variability. Type III mixed-effects ANOVA with Wald χ² tests was used to show the main effect of each independent variable. For significant omnibus effects, pairwise comparisons were conducted to identify specific differences between stimulation conditions, with Tukey’s correction applied to control for multiple comparisons. Models were first estimated with both main effects and the Location × Frequency interaction term; because the interaction was not statistically significant across outcomes, final models included only the main effects. Multilevel models were estimated in R (RStudio) [54, 55] using *lme4* [56]; Type III Wald χ² tests were conducted with the *car* package [57], and Tukey-adjusted pairwise contrasts were obtained via *emmeans* [58].

We conducted a secondary analysis to calculate the proportion of participants that could tolerate stimulation at clinically relevant levels of 100% sMT and 120% sMT. We used a generalized estimating equations (GEE) analyses with a binomial distribution and logit link to assess tolerability as a function of stimulation location and frequency, computed in IBM SPSS Statistics version 29 (IBM Corp., Armonk, NY). An independent working correlation structure was used.

## RESULTS

### Participant Characterization

Of 20 individuals who consented to participate in the study, 18 were able to complete the procedures and included in the analysis (1 dropout due to scheduling, 1 was unable to complete due to discomfort).

The community sample had a broad age range from 18 – 82 years of age, with mean age of 34.8 years (*SD* = 21.6 years). There were 12 female and 6 male participants. Symptoms of depression were overall minimal to mild (PHQ-9: range 0-16, mean = 2.8, *SD* = 4.4), as were symptoms of anxiety (GAD-7: range 0-19, mean = 2.8, *SD* = 4.6). Most participants had no-to-little recent pain of significant impact (PROMIS Pain Interference Short Form 6b: range 6-18, mean = 7.8, *SD* = 3.4).

The mean scalp distance from Beam F3 to THREE-D as measured on a marked headcap was 3.7 cm (*SD* = 0.8 cm, min = 1.4, max = 5.1). The mean scalp MNI coordinates for Beam F3 were -50.4, 28.2, 56.6 (*SD* x= 4.4, y = 6.3, z = 4.3), which we estimate correspond to a cortical location of -44, 22, 51. We estimate the scalp location of the THREE-D target as -44, 50, 32. The direct line distance between estimated scalp locations is 3.3 cm, compared to the measured 3.7 cm along the curved scalp.

The average MT for participants was 48.5% of maximum machine output (range 36-69, *SD* = 9.0). This study average MT is greater than the established system MT of 39% maximum machine output, so the converted study average is 124% sMT (range 92-177, *SD* = 23). This would indicate that the study average MT was 124% higher than that broader population average.

### Sensory Thresholds

The overall mean sensory threshold was 40.2% sMT (*SD* = 9.4). Table 1a provides the mean sensory thresholds for each location-frequency combination, as well as cumulative means for each location and each frequency.

**Table 1a.** Sensory Threshold Descriptive Statistics. Mean % sMT of Sensory Thresholds (±SD)

|  | 1 Hz |  | 10 Hz |  | iTBS |  | Total |  |
| --- | --- | --- | --- | --- | --- | --- | --- | --- |
| | Mean | ±SD | Mean | ±SD | Mean | ±SD | Mean | ±SD |
| **Knee** | 45.3 | 8.6 | 38.5 | 8.6 | 39.0 | 7.4 | 40.9 | 8.7 |
| **BeamF3** | 41.3 | 7.7 | 38.5 | 9.3 | 35.0 | 7.9 | 38.3 | 8.6 |
| **THREE-D** | 38.5 | 7.0 | 32.8 | 6.3 | 32.2 | 6.2 | 34.5 | 7.0 |
| **AF8** | 39.0 | 8.2 | 36.8 | 9.5 | 35.6 | 9.8 | 37.1 | 9.1 |
| **mPFC** | 48.1 | 10.2 | 43.0 | 10.7 | 45.3 | 11.7 | 45.5 | 10.9 |
| **M1** | 43.0 | 8.8 | 41.3 | 9.2 | 41.3 | 9.2 | 41.9 | 8.9 |
| **P3** | 46.4 | 10.3 | 40.7 | 5.6 | 41.9 | 6.1 | 43.0 | 7.9 |
| **Total** | 43.1 | 9.2 | 38.8 | 9.0 | 38.6 | 9.4 | 40.2 | 9.4 |

Multilevel modeling indicated significant main effects of stimulation location and frequency on sensory threshold values. Tukey-adjusted pairwise comparisons are shown in Table 1b. Stimulation location significantly influenced sensory thresholds, χ²(6) = 88.41, *p* < .001. Compared with AF8, higher stimulation intensity was required for sensory detection at M1 (*d* = −0.66, *p* = .012), mPFC (*d* = −1.16, *p* < .001), and P3 (*d* = −0.82, *p* = .001). THREE-D sensory detection occurred at lower intensity relative to Knee (*d* = 0.90, *p* < .001), M1 (*d* = 1.03, *p* < .001), mPFC (*d* = 1.53, *p* < .001), and P3 (*d* = 1.19, *p* < .001). Beam F3 sensory detection also occurred at lower intensity relative than mPFC (*d* = −1.00, *p* < .001) and P3 (*d* = −0.66, *p* = .012). No other pairwise comparisons were statistically significant (*ds* ≤ 0.53, *ps* ≥ .092).

**Table 1b.** Tukey-Adjusted Pairwise Comparisons for Sensory Thresholds by <u>Stimulation Frequency and Location</u>.

| contrast | estimate | SE | df | CI | p value | d |
| --- | --- | --- | --- | --- | --- | --- |
| <u>FREQUENCY</u> |  |  |  |  |  |  |
| **1 Hz - 10 Hz** | **4.31** | **0.91** | **352** | **[2.17, 6.45]** | **< .001** | **0.6** |
| **1 Hz - iTBS** | **4.48** | **0.91** | **352** | **[2.34, 6.62]** | **< .001** | **0.62** |
| 10 Hz - iTBS | 0.16 | 0.91 | 352 | [-1.98, 2.3] | 0.982 | 0.02 |
| <u>LOCATION</u> |  |  |  |  |  |  |
| AF8 - BeamF3 | -1.14 | 1.39 | 352 | [-5.26, 2.98] | 0.983 | -0.16 |
| AF8 - Knee | -3.8 | 1.39 | 352 | [-7.92, 0.32] | 0.092 | -0.53 |
| **AF8 - M1** | **-4.75** | **1.39** | **352** | **[-8.87, -0.63]** | **0.012** | **-0.66** |
| **AF8 - mPFC** | **-8.36** | **1.39** | **352** | **[-12.47, -4.24]** | **< .001** | **-1.16** |
| **AF8 - P3** | **-5.89** | **1.39** | **352** | **[-10.01, -1.77]** | **0.001** | **-0.82** |
| AF8 - (THREE-D) | 2.66 | 1.39 | 352 | [-1.46, 6.78] | 0.472 | 0.37 |
| BeamF3 - Knee | -2.66 | 1.39 | 352 | [-6.78, 1.46] | 0.472 | -0.37 |
| BeamF3 - M1 | -3.61 | 1.39 | 352 | [-7.73, 0.51] | 0.129 | -0.5 |
| **BeamF3 - mPFC** | **-7.22** | **1.39** | **352** | **[-11.33, -3.1]** | **< .001** | **-1** |
| **BeamF3 - P3** | **-4.75** | **1.39** | **352** | **[-8.87, -0.63]** | **0.012** | **-0.66** |
| BeamF3 - (THREE-D) | 3.8 | 1.39 | 352 | [-0.32, 7.92] | 0.092 | 0.53 |
| Knee - M1 | -0.95 | 1.39 | 352 | [-5.07, 3.17] | 0.993 | -0.13 |
| **Knee - mPFC** | **-4.56** | **1.39** | **352** | **[-8.68, -0.44]** | **0.019** | **-0.63** |
| Knee - P3 | -2.09 | 1.39 | 352 | [-6.21, 2.03] | 0.742 | -0.29 |
| **Knee - (THREE-D)** | **6.46** | **1.39** | **352** | **[2.34, 10.57]** | **< .001** | **0.9** |
| M1 - mPFC | -3.61 | 1.39 | 352 | [-7.73, 0.51] | 0.129 | -0.5 |
| M1 - P3 | -1.14 | 1.39 | 352 | [-5.26, 2.98] | 0.983 | -0.16 |
| **M1 - (THREE-D)** | **7.41** | **1.39** | **352** | **[3.29, 11.52]** | **< .001** | **1.03** |
| mPFC - P3 | 2.47 | 1.39 | 352 | [-1.65, 6.59] | 0.564 | 0.34 |
| **mPFC - (THREE-D)** | **11.02** | **1.39** | **352** | **[6.9, 15.13]** | **< .001** | **1.53** |
| **P3 - (THREE-D)** | **8.55** | **1.39** | **352** | **[4.43, 12.66]** | **< .001** | **1.19** |

**Note.** Results reflect Tukey-adjusted post-hoc pairwise comparisons from multilevel models. Positive estimates indicate that the first condition listed in the contrast required a higher stimulation intensity to reach the sensory threshold than the second. *SE* = standard error; *df* = degrees of freedom; *CI* = 95% confidence interval; *d* = Cohen's *d*. Statistically significant comparisons are shown in bold.

Frequency significantly also predicted sensory thresholds, χ²(2) = 31.22, *p* < .001. Tukey-adjusted pairwise comparisons (Table 1b) showed that 1 Hz stimulation required higher intensity for sensory detection than both 10 Hz (*d* = 0.60, *p* < .001) and iTBS (*d* = 0.62, *p* < .001), whereas 10 Hz and iTBS did not differ (*p* = .98).

### Annoyance Thresholds

In 15.9% of all trials (60 of 378), stimulation was stopped at or near the participant’s 120% motor-threshold safety limit before the annoyance threshold was reached. In these cases, the stopping value was treated as the annoyance threshold in analyses. Across locations, early stops were most common at the knee (15 of 60), P3 (15 of 60), M1 (12 of 60), and mPFC (9 of 60) sites. Early stops occurred most frequently during 1 Hz stimulation (36 of 60). Accordingly, annoyance thresholds may be modestly underestimated, particularly for 1 Hz stimulation.

The overall mean annoyance threshold was 87.6% sMT (*SD* = 29.1). Table 2a provides the mean annoyance thresholds for each location-frequency combination, as well as cumulative means for each location and each frequency.

**Table 2a.** Annoyance Threshold Descriptive Statistics. Mean % sMT of Annoyance Thresholds (±SD)

|  | 1 Hz |  | 10 Hz |  | iTBS |  | Total |  |
| --- | --- | --- | --- | --- | --- | --- | --- | --- |
| | Mean | ±SD | Mean | ±SD | Mean | ±SD | Mean | ±SD |
| **Knee** | 114.8 | 25.9 | 98.3 | 26.8 | 85.8 | 27.1 | 99.6 | 28.7 |
| **BeamF3** | 96.6 | 27.0 | 80.6 | 23.3 | 73.2 | 19.0 | 83.5 | 24.9 |
| **THREE-D** | 86.9 | 24.3 | 67.0 | 20.4 | 59.5 | 19.4 | 71.1 | 24.1 |
| **AF8** | 72.6 | 27.7 | 61.8 | 24.1 | 55.6 | 22.0 | 63.3 | 25.2 |
| **mPFC** | 112.0 | 22.3 | 89.7 | 23.1 | 88.0 | 25.4 | 96.6 | 25.7 |
| **M1** | 105.1 | 23.7 | 95.4 | 26.6 | 87.5 | 24.5 | 96.0 | 25.5 |
| **P3** | 114.8 | 22.6 | 98.9 | 23.3 | 96.0 | 24.2 | 103.2 | 24.4 |
| **Total** | 100.4 | 28.5 | 84.5 | 27.4 | 77.9 | 26.9 | 87.6 | 29.1 |

Multilevel modeling indicated significant effects of stimulation location and frequency on annoyance thresholds. Tukey-adjusted pairwise comparisons are shown in Table 2b. Stimulation location significantly influenced annoyance thresholds, χ²(6) = 296.48, *p* < .001. AF8 had a substantially lower annoyance threshold than all other sites except THREE-D, including Beam F3 (*d* = −1.25, *p* < .001), Knee (*d* = −2.26, *p* < .001), M1 (*d* = −2.03, *p* < .001), mPFC (*d* = −2.07, *p* < .001), and P3 (*d* = −2.48, *p* < .001). THREE-D stimulation had a lower annoyance threshold than several other locations including Beam F3 (*d* = 0.77, *p* = .002), Knee (*d* = 1.77, *p* < .001), M1 (*d* = 1.55, *p* < .001), mPFC (*d* = 1.58, *p* < .001), and P3 (*d* = 2.00, *p* < .001). Finally, Beam F3 had a lower annoyance threshold than Knee (*d* = −1.00, *p* < .001), M1 (*d* = −0.78, *p* = .001), mPFC (*d* = −0.82, *p* = .001), and P3 (*d* = −1.23, *p* < .001). No other pairwise differences were significant (*ds* ≤ 0.48, *ps* ≥ .156).

**Table 2b.** Tukey-Adjusted Pairwise Comparisons for Annoyance Thresholds by Stimulation Frequency and Location.

| contrast | estimate | SE | df | CI | p value | d |
| --- | --- | --- | --- | --- | --- | --- |
| <u>FREQUENCY</u> |  |  |  |  |  |  |
| **1 Hz - 10 Hz** | **15.87** | **2.02** | **352** | **[11.11, 20.64]** | **< .001** | **0.99** |
| **1 Hz - iTBS** | **22.47** | **2.02** | **352** | **[17.7, 27.23]** | **< .001** | **1.4** |
| **10 Hz - iTBS** | **6.59** | **2.02** | **352** | **[1.83, 11.36]** | **0.004** | **0.41** |
| <u>LOCATION</u> |  |  |  |  |  |  |
| **AF8 - BeamF3** | **-20.13** | **3.09** | **352** | **[-29.3, -10.97]** | **< .001** | **-1.25** |
| **AF8 - Knee** | **-36.28** | **3.09** | **352** | **[-45.45, -27.11]** | **< .001** | **-2.26** |
| **AF8 - M1** | **-32.67** | **3.09** | **352** | **[-41.84, -23.5]** | **< .001** | **-2.03** |
| **AF8 - mPFC** | **-33.24** | **3.09** | **352** | **[-42.41, -24.07]** | **< .001** | **-2.07** |
| **AF8 - P3** | **-39.89** | **3.09** | **352** | **[-49.05, -30.72]** | **< .001** | **-2.48** |
| AF8 - (THREE-D) | -7.79 | 3.09 | 352 | [-16.96, 1.38] | 0.156 | -0.48 |
| **BeamF3 - Knee** | **-16.14** | **3.09** | **352** | **[-25.31, -6.98]** | **< .001** | **-1** |
| **BeamF3 - M1** | **-12.54** | **3.09** | **352** | **[-21.7, -3.37]** | **0.001** | **-0.78** |
| **BeamF3 - mPFC** | **-13.11** | **3.09** | **352** | **[-22.27, -3.94]** | **0.001** | **-0.82** |
| **BeamF3 - P3** | **-19.75** | **3.09** | **352** | **[-28.92, -10.59]** | **< .001** | **-1.23** |
| **BeamF3 - (THREE-D)** | **12.35** | **3.09** | **352** | **[3.18, 21.51]** | **0.002** | **0.77** |
| Knee - M1 | 3.61 | 3.09 | 352 | [-5.56, 12.78] | 0.906 | 0.22 |
| Knee - mPFC | 3.04 | 3.09 | 352 | [-6.13, 12.21] | 0.957 | 0.19 |
| Knee - P3 | -3.61 | 3.09 | 352 | [-12.78, 5.56] | 0.906 | -0.22 |
| **Knee - (THREE-D)** | **28.49** | **3.09** | **352** | **[19.32, 37.66]** | **< .001** | **1.77** |
| M1 - mPFC | -0.57 | 3.09 | 352 | [-9.74, 8.6] | 1 | -0.04 |
| M1 - P3 | -7.22 | 3.09 | 352 | [-16.39, 1.95] | 0.231 | -0.45 |
| **M1 - (THREE-D)** | **24.88** | **3.09** | **352** | **[15.71, 34.05]** | **< .001** | **1.55** |
| mPFC - P3 | -6.65 | 3.09 | 352 | [-15.82, 2.52] | 0.326 | -0.41 |
| **mPFC - (THREE-D)** | **25.45** | **3.09** | **352** | **[16.28, 34.62]** | **< .001** | **1.58** |
| **P3 - (THREE-D)** | **32.1** | **3.09** | **352** | **[22.93, 41.27]** | **< .001** | **2** |

**Note.** Results reflect Tukey-adjusted post-hoc pairwise comparisons from multilevel models. Positive estimates indicate that the first condition listed in the contrast required a higher stimulation intensity to reach the annoyance threshold than the second. *SE* = standard error; *df* = degrees of freedom; *CI* = 95% confidence interval; *d* = Cohen's *d*. Statistically significant comparisons are shown in bold.

Frequency also significantly predicted the stimulation intensity required to reach annoyance, χ²(2) = 130.23, *p* < .001. Tukey-adjusted pairwise comparisons (Table 2b) showed that 1 Hz stimulation had a higher annoyance threshold than 10 Hz (*d* = 0.99, *p* < .001) and iTBS (*d* = 1.40, *p* < .001). Additionally, 10 Hz stimulation had a higher annoyance threshold than iTBS (*d* = 0.41, *p* = .004).

### Tolerability Thresholds

In 39.4% of all trials (149 of 378), stimulation was stopped at or near the participant’s 120% motor-threshold safety limit before reaching the tolerability threshold. In these cases, the stopping value was treated as the tolerability threshold in analyses. As a result, group means likely underestimate true tolerability thresholds, particularly for conditions in which participants reached the safety ceiling before reporting intolerability.

The overall mean tolerability threshold was 111.5% sMT (*SD* = 28.5). Table 3a provides the mean tolerability thresholds for each location-frequency combination, as well as cumulative means for each location and each frequency.

**Table 3a.** Tolerability Threshold Descriptive Statistics. Mean % sMT of Tolerability Thresholds (±SD)

|  | 1 Hz |  | 10 Hz |  | iTBS |  | Total |  |
| --- | --- | --- | --- | --- | --- | --- | --- | --- |
| | Mean | ±SD | Mean | ±SD | Mean | ±SD | Mean | ±SD |
| **Knee** | 136.5 | 21.5 | 125.1 | 19.7 | 121.1 | 24.5 | 127.5 | 22.5 |
| **BeamF3** | 116.0 | 22.3 | 104.6 | 23.7 | 96.6 | 22.2 | 105.7 | 23.7 |
| **THREE-D** | 112.5 | 28.5 | 89.7 | 23.6 | 79.5 | 22.8 | 93.9 | 28.3 |
| **AF8** | 102.8 | 25.6 | 79.5 | 28.8 | 73.2 | 25.9 | 85.2 | 29.3 |
| **mPFC** | 133.0 | 18.5 | 114.8 | 19.7 | 110.8 | 22.4 | 119.6 | 22.1 |
| **M1** | 129.1 | 22.3 | 116.5 | 22.3 | 117.1 | 19.3 | 120.9 | 21.7 |
| **P3** | 137.6 | 20.5 | 124.5 | 14.6 | 121.7 | 14.9 | 127.9 | 18.0 |
| **Total** | 123.9 | 25.7 | 107.8 | 27.0 | 102.8 | 28.4 | 111.5 | 28.5 |

Multilevel modeling indicated significant main effects of stimulation location and frequency on tolerability threshold values. Tukey-adjusted pairwise comparisons are shown in Table 3b. Stimulation location significantly influenced tolerability thresholds, χ²(6) = 279.51, *p* < .001. Relative to AF8, the tolerability threshold was higher at Knee (*d* = −2.33, *p* < .001), P3 (*d* = −2.35, *p* < .001), M1 (*d* = −1.96, *p* < .001), mPFC (*d* = −1.89, *p* < .001), and BeamF3 (*d* = −1.13, *p* < .001). BeamF3 had a tolerability threshold lower than Knee (*d* = −1.20, *p* < .001), P3 (*d* = −1.22, *p* < .001), M1 (*d* = −0.83, *p* < .001), and mPFC (*d* = −0.76, *p* = .002). THREE-D had a tolerability threshold lower than Knee (*d* = 1.85, *p* < .001), P3 (*d* = 1.87, *p* < .001), M1 (*d* = 1.48, *p* < .001), mPFC (*d* = 1.41, *p* < .001), and BeamF3 (*d* = 0.65, *p* = .015). No other pairwise differences were statistically significant (*ds* ≤ 0.48, *ps* ≥ .165).

**Table 3b.**
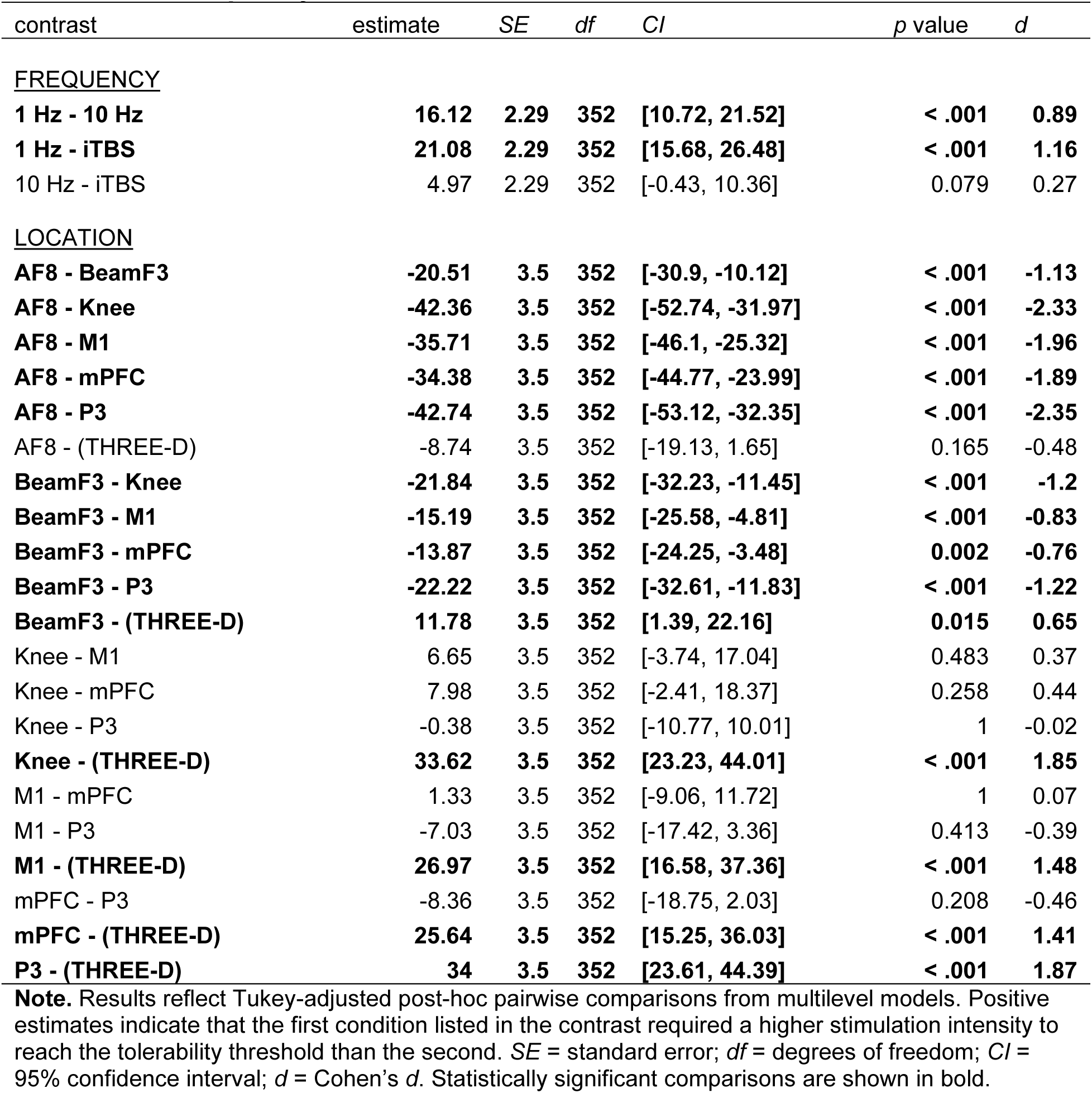
Tukey-Adjusted Pairwise Comparisons for Tolerability Thresholds by <u>Stimulation Frequency and Location</u>.

Frequency also significantly predicted tolerability thresholds, χ²(2) = 92.38, *p* < .001. Tukey-adjusted pairwise comparisons (Table 3b) showed that 1 Hz stimulation had a higher tolerability threshold than 10 Hz (*d* = 0.89, *p* < .001) and iTBS (*d* = 1.16, *p* < .001). The threshold difference between 10 Hz and iTBS was marginally significant (*d* = 0.27, *p* = .079).

### Relation of Thresholds

The overall mean sensory threshold was 40.2% sMT (*SD* = 9.4), the overall mean annoyance threshold was 87.6% sMT (*SD* = 29.1), and the overall mean tolerability threshold was 111.5% sMT (*SD* = 28.5). Figure 1 portrays the relationships of thresholds, by location and frequency.

**Figure 1.**
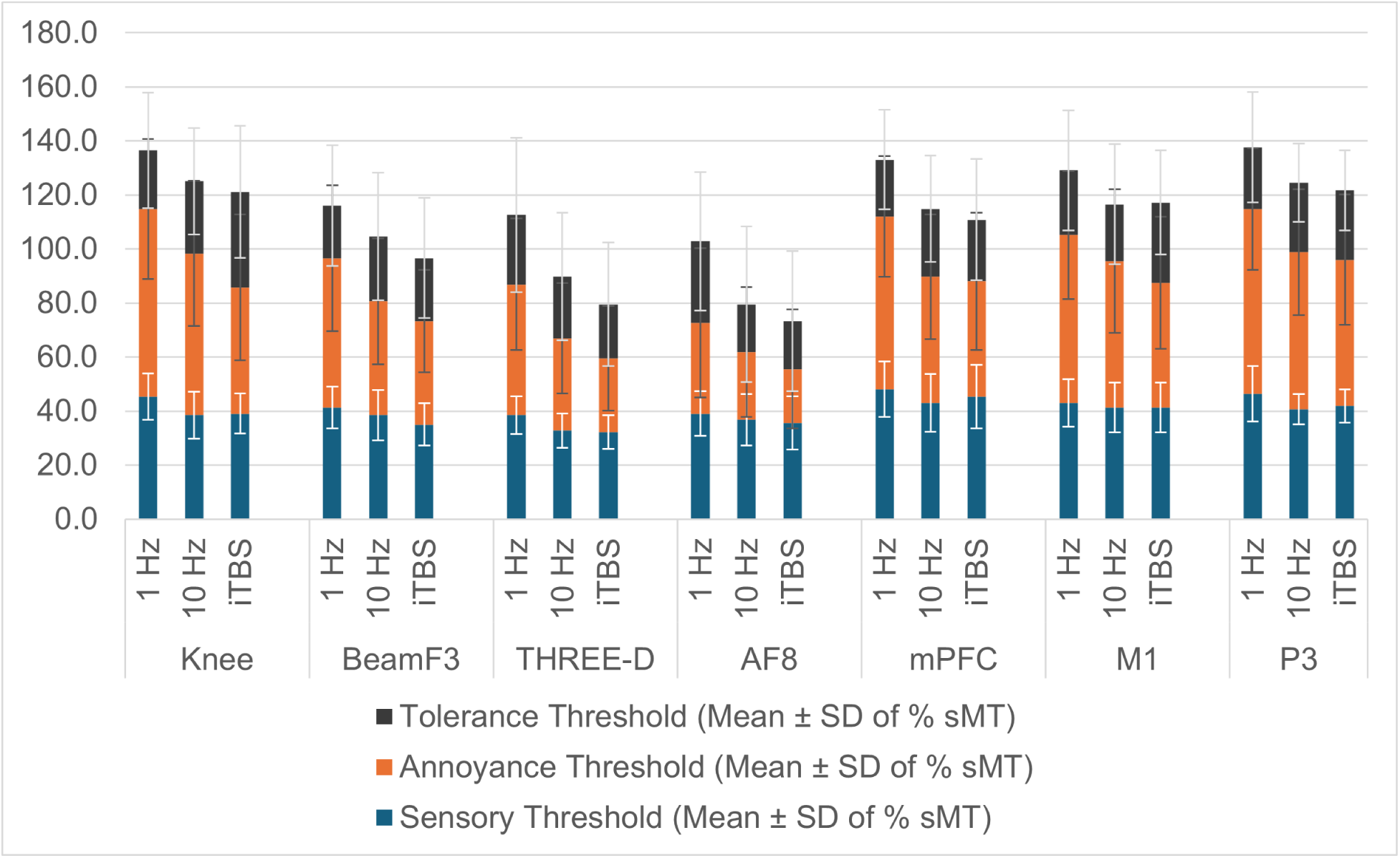
Relationships of Thresholds, by Location and Frequency.

### Tolerability at 100% sMT and 120% sMT

Given that all participants could safely be tested at 100% sMT (n=18) and most (n=16) participants could be safely tested at 120% sMT, we are able to calculate the proportion of participants that could tolerate stimulation at these relevant levels. The percentage of participants that could tolerate stimulation at each of the 21 trials (7 locations x 3 frequencies) are calculated for 100% sMT (Figure 2a) and 120% sMT (Figure 2b).

**Figure 2.**
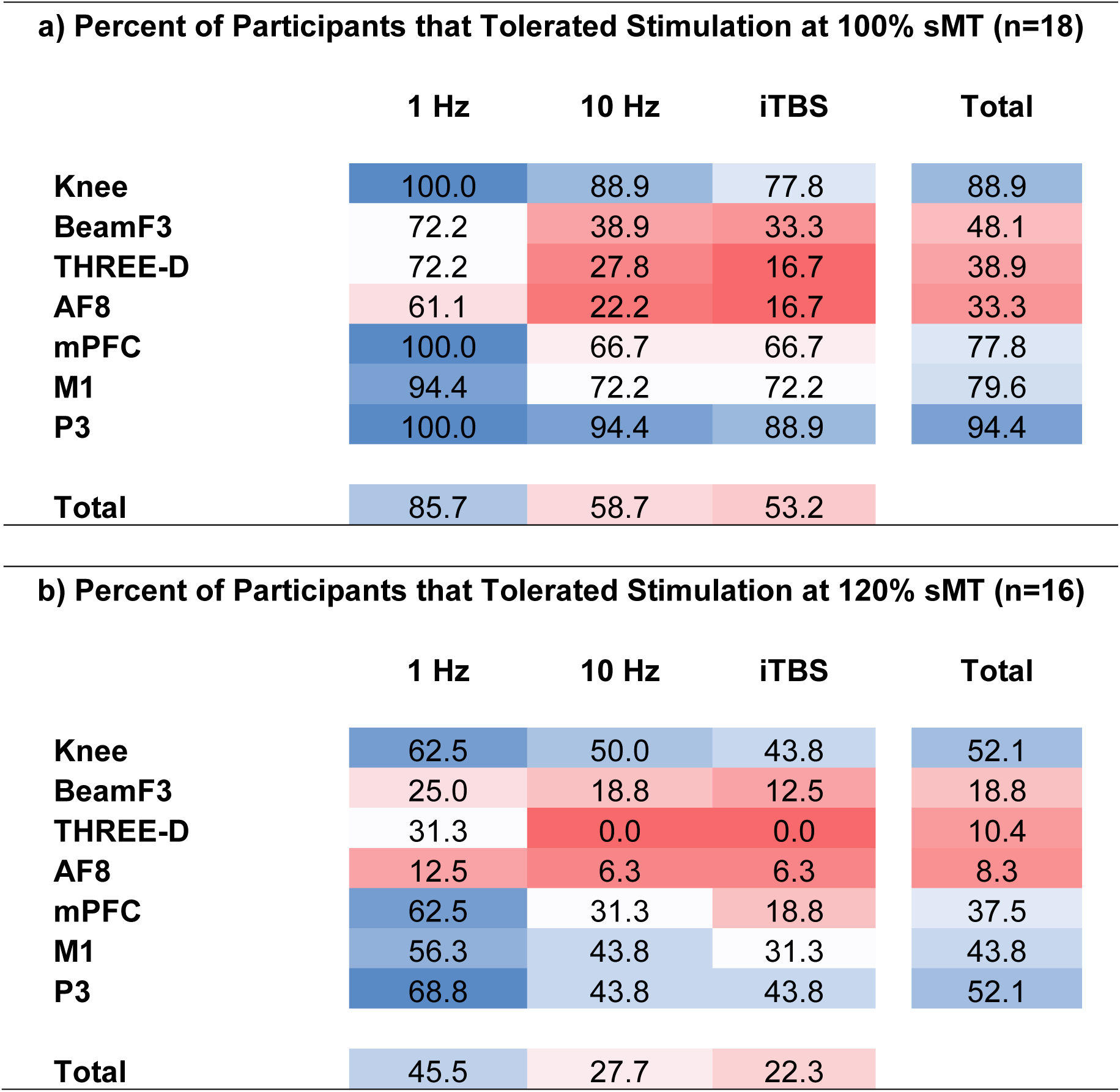
Percent of Participants Tolerating Stimulation at Common Treatment Levels.

At 100% sMT, we found significant main effects of both location, Wald *χ^2^*(6) = 71.60, p < .001, and frequency, Wald *χ^2^* (2) = 27.86, p < .001. For location, estimated marginal means (proportion of tolerated trials) indicated that stimulation was most tolerable at P3 (mean = .96, *SE* = .03) and Knee (mean = .92, *SE* = .04), followed by M1 (mean = .84, *SE* = .07) and mPFC (mean = .82, *SE* = .06). Stimulation was less tolerated at Beam F3 (mean = .49, *SE* = .12), THREE-D (mean = .37, *SE* = .09), and AF8 (mean = .31, *SE* = .10). Pairwise comparisons showed that P3, Knee, M1, and mPFC each were statistically significantly more tolerable than Beam F3, THREE-D, and AF8 (all Bonferroni-adjusted *p* values < .05). No other comparisons had statistically significant differences.

For frequency, 1 Hz stimulation was the most tolerable (mean = .92, *SE* = .03), compared to 10 Hz (mean = .63, *SE* = .09) and iTBS (mean = .55, *SE* = .09). Pairwise comparisons showed that 1 Hz was statistically significantly more tolerable than both 10 Hz and iTBS (Bonferroni-adjusted *p* values < .001). The difference between 10 Hz and iTBS was not statistically significant (*p* = .149).

At 120% sMT, we found significant main effects of both location, Wald *χ^2^*(6) = 32.55, p < .001, and frequency, Wald *χ^2^* (2) = 18.73, p < .001. For location, estimated marginal means (proportion of tolerated trials) indicated that stimulation was most tolerable at P3 (mean = .52, *SE* = .11) and Knee (mean = .52, *SE* = .12), followed by M1 (mean = .43, *SE* = .12) and mPFC (mean = .37, *SE* = .10). Stimulation was less tolerated at Beam F3 (mean = .17, *SE* = .09), THREE-D (mean = .09, *SE* = .04), and AF8 (mean = .07, *SE* = .06). Pairwise comparisons showed that P3 and Knee were statistically significantly more tolerable than Beam F3, THREE-D, and AF8 (all Bonferroni-adjusted *p* values < .05), while M1 and mPFC were statistically significantly more tolerable than just THREE-D and AF8 (all Bonferroni-adjusted *p* values < .05). No other comparisons had statistically significant differences.

For frequency, 1 Hz stimulation was the most tolerable (mean = .44, *SE* = .11), compared to 10 Hz (mean = .23, *SE* = .08) and iTBS (mean = .18, *SE* = .06). Pairwise comparisons showed that 1 Hz was statistically significantly more tolerable than both 10 Hz and iTBS (Bonferroni-adjusted *p* values < .01). The difference between 10 Hz and iTBS was not statistically significant (*p* = .144).

### Tolerability Survival Curves

To visually characterize tolerability, we graphed the percentage of participants that tolerated a stimulation trial (y-axis) at each increasing level of machine output (x-axis). Figure 3a shows the average percent tolerability by location, aligning with the statistical analysis showing the best tolerability for P3 and Knee, followed by closely by M1, and mPFC. Beam F3, THREE-D, and AF8 were less tolerable, with more participants rating “troublesome or miserable” at lower TMS machine output. Figure 3b shows the average percent tolerability by frequency, aligning with the statistical analysis showing the best tolerability for 1 Hz.

**Figure 3.**
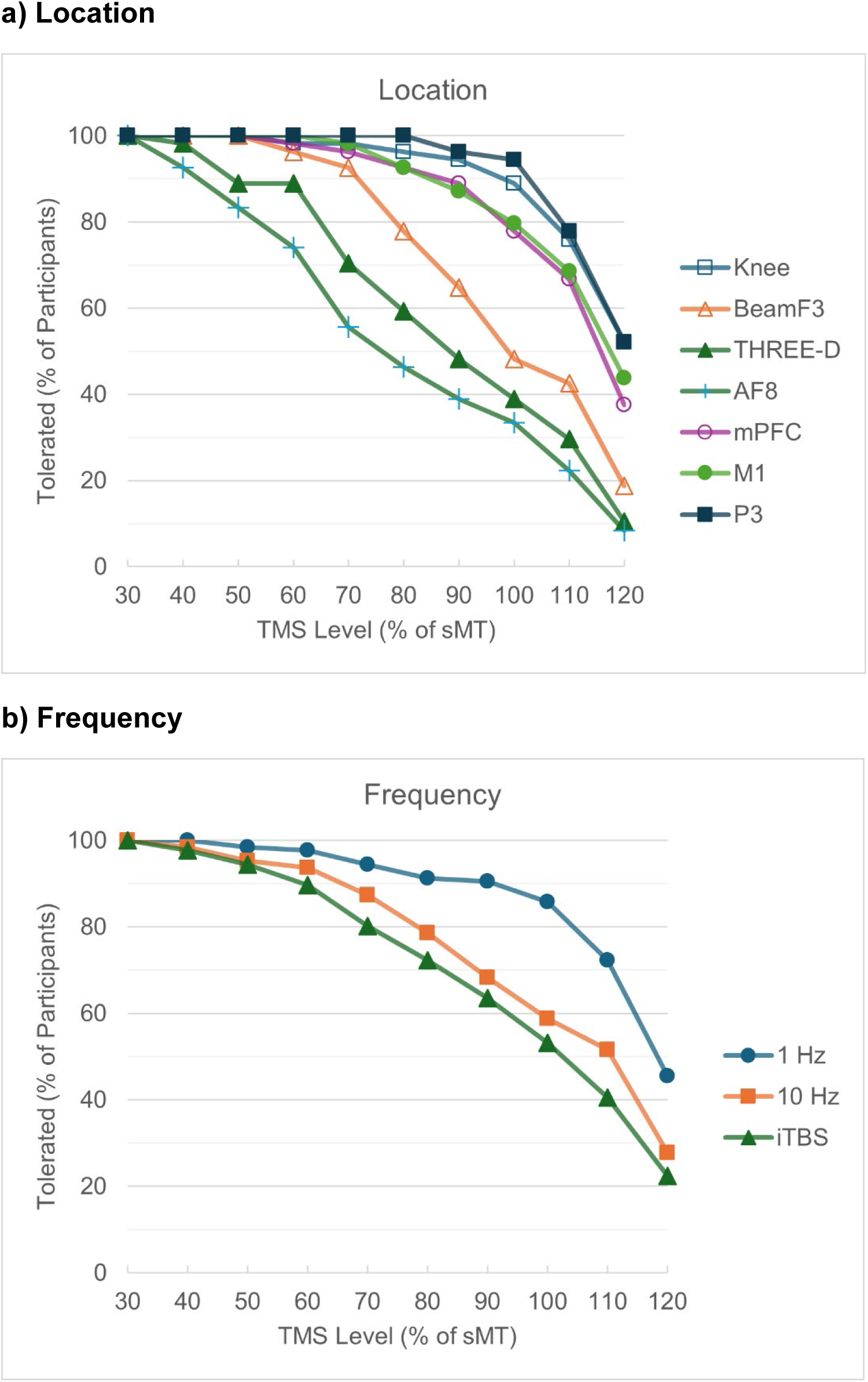
Survival Curves for Tolerability by Location and Frequency.

### Verbal Descriptors

As an exploratory qualitative analysis, we calculated the most frequently selected terms (mode) on the McGill pain questionnaire, describing the sensation at the end of each trial. Importantly, many items were often “none/not applicable”. As shown in Table 4, there were similarities with frontal locations (Beam F3, THREE-D, and AF8) for items 1, 4, 16, and 17 (indicating throbbing, sharp, troublesome, and penetrating/piercing sensation) differentiated from other locations. With frequency, 10 Hz and iTBS were similar for items 4, 8, 16, 17 (sharp, tingling, troublesome and penetrating sensation) differentiated from 1 Hz.

**Table 4.** McGill Pain Questionnaire Descriptors (Mode)

|  | **McGill-1** | **McGill-2** | **McGill-4** | **McGill-8** | **McGill-16** | **McGill-17** |
| --- | --- | --- | --- | --- | --- | --- |
| **Knee** | Flickering | Jumping | N/A | Tingling | N/A | N/A |
| **BeamF3** | Throbbing | Jumping | Sharp | N/A | Troublesome | Penetrating |
| **THREE-D** | Throbbing | Shooting | Sharp | Tingling | Troublesome | Piercing |
| **AF8** | Throbbing | Shooting | Sharp | N/A | Troublesome | Penetrating |
| **mPFC** | Beating | Jumping | Sharp | N/A | Troublesome | N/A |
| **M1** | Flickering | Jumping | N/A | Tingling | Annoying | N/A |
| **P3** | Pulsing | Shooting | N/A | N/A | Annoying | N/A |
| **1 Hz** | Flickering | Jumping | N/A | N/A | Annoying | N/A |
| **10 Hz** | Flickering | Shooting | Sharp | Tingling | Troublesome | Penetrating |
| **iTBS** | Pulsing | Jumping | Sharp | Tingling | Troublesome | Penetrating |

## DISCUSSION

This study was motivated by the lack of within-individual data comparing TMS scalp sensations across both multiple locations and frequencies, with a focus on parameter combinations that are relevant to existing and emerging clinical applications. Although TMS researchers and clinicians have developed experiential knowledge about tolerability, the sensory experience of TMS has not been systematically quantified. Tactile sensations may have an underappreciated impact on research outcomes, may interact with brain state and response to TMS, and can constrain the range of stimulation parameters that are feasible due to tolerability.

The location of stimulation has a significant impact on the tolerability of TMS. Knowledge in this regard is not well-established, with general theory supporting locations with greater cranial nerve and muscle involvement as plausibly less comfortable for stimulation. Our results support this theory, with prefrontal sites found to be less tolerable than others tested. The least comfortable and tolerable stimulation sites were AF8 and THREE-D close to facial muscles and nerves, followed by Beam F3, followed by mPFC, M1, P3, and Knee with similar levels as most tolerable. We did find support for our hypothesis that TMS is less tolerable at the THREE-D prefrontal and orbitofrontal locations relative to the modified Beam F3 prefrontal location. Our results generally concur with an extensive study mapping the effects of 43 sites with single-pulse stimulation in the context of task performance [2]. However, our study tested multiple frequencies and locations relevant to clinical interventions. We limited the number of sites tested considering participant burden, as well as only tested one coil orientation per site. Research shows that coil orientation can impact tolerability [59], and our experience suggests the small adjustments in scalp location and coil angle can substantially improve patient comfort (especially if the coil is directly over a cranial nerve).

The frequency of stimulation also has a significant impact on the tolerability of TMS. Clinicians generally consider 1 Hz stimulation to be more tolerable than 10 Hz [60]. Our results clearly support this understanding both in thresholds and qualitative ratings (MPQ), aligning with a similar study testing 1 Hz and 10 Hz at three locations [9]. The tolerability difference between 10 Hz and iTBS is less clear in the TMS field, although our experience and a secondary analysis of the THREE-D study suggest that 10 Hz may be slightly less painful than iTBS [14]. Our results suggest that while iTBS is a more annoying sensation than 10 Hz stimulation, the tolerability at therapeutic levels are marginally different in thresholds and qualitative ratings (MPQ). We did not find strong support for our hypothesis that TMS theta burst is less tolerable than 10 Hz, although we did not test at durations and intensities equivalent to therapeutic sessions (e.g. iTBS for 3 minutes or greater, at 120% MT) [36]. To our knowledge, this is the first study to directly compare 1 Hz, 10 Hz and iTBS within-individuals. We did not test 5 Hz considering participant burden. However, some clinicians consider 5 Hz as an alternative to 10 Hz, when 10 Hz is not tolerable [61]. Novel frequency patterns that produce desired brain effects, while improving the sensory experience, is a potential opportunity for future development.

We characterized the overall mean sensory threshold as 40.2% sMT with a relatively low amount of variability. The sensory threshold provides a clear indication of biological effect in the superficial tissue, analogous to the motor threshold that provides a clear indication of biological effects in the brain. Peripheral magnetic stimulation may be used to treat pain [62, 63]. Sensory thresholds, along with other thresholds and qualitative data, can inform this application. Peripheral stimulation of cranial nerves is emerging for psychiatric conditions [31, 32], and reports of low intensity TMS may have relatively more sensory effects than brain effects [64]. In clinical research of TMS, the peripheral effects have largely been considered a nuisance variable, when in fact it may contribute to therapeutic benefit. Conversely, the brain effects of TMS may also impact the perception of pain from TMS [41].

We characterized the overall mean annoyance threshold as 87.6% sMT with considerable variability. The annoyance threshold likely has the greatest relevance for studies where attention and other cognitive processes are important. We infer that annoying sensory stimulation represents a level that starts to distract from other sensory and cognitive processes. We found effects of both location and frequency on the annoyance threshold, so studies comparing location and frequency, especially above 90% sMT are likely to be confounded by tactile components of TMS.

Several limitations of this study warrant consideration. First, some testing was terminated early due to safety guidelines. As a result, group means likely underestimate true tolerability thresholds, particularly for conditions in which participants reached the safety ceiling before reporting intolerability. Second, we only tested two trains before adjusting to a higher intensity. Sensory experience may change over time and many TMS protocols (especially interventions) stimulate for much longer periods. Within a session repeated stimulation could become more tolerable with adaption or more troublesome with sensitization. A caveat of our methodology is that we defined tolerability from the McGill Pain scale anchors of “troublesome or miserable” on an item that extends up to “unbearable”. We selected the lower valence as more relevant for tolerability in real-world situations. The conceptualization of tolerance can vary based on perceived benefits or appraisal of harm. For instance, the motivation and ability to tolerate discomfort may be lower for an experimental research study in healthy participants relative to an individual receiving TMS to treat a disabling condition. Our population sample was relatively healthy without a treatment incentive for participating. Of note, our population had a higher MT (48.5% of maximum machine output) than the manufacturer provided MT (39%) used to calculate sMT, and we have also seen a higher MT (46.9%) in a separate study on mental health conditions on the same device [65]. This does not impact the overall relative conclusions but would impact interpretation of values generalized to other populations. We only tested one coil type, and sensations may vary with the broader range of field characteristics [5, 6].

An overarching objective of the project was to advance QST methods for measuring TMS tactile sensations and tolerability, as well as to highlight the underappreciated impact of the TMS sensory component. Future work should test tolerability over longer periods of time, compare coil types, and explore methods to increase tolerability and comfort for clinical applications [40, 66, 67]. We tested the knee as a surrogate for more extensive testing that may exceed safety considerations for direct brain stimulation. Our data suggests that the knee is a good representative for posterior scalp regions, but less so for frontal regions. Future research may explore whether there is another location (e.g., hand) that has a better profile to model frontal scalp sensations.

Our qualitative analysis used standardized wording that showed some alignment with threshold testing, however additional work using unconstrained descriptors may be beneficial. The qualitative aspects may also point to specific nerve fibers impacted. Sharp, tingling pain may link to fast-conducting A-delta fiber activation, whereas dull or slower throbbing pain may link to slow C-fiber mediated nociception [68]. An important future direction is to better understand the contribution of sensory stimulation as a component of therapeutic benefit. The appropriate sham condition has been a challenge for TMS researchers, with examples of trials where both active and sham demonstrated expected effects. While placebo effects and behavioral activation may account for some therapeutic benefit in sham arms, it is plausible that that peripheral stimulation contributes as well.

This study quantifies the impact of location and frequency, with increasing stimulation intensity, on the sensation and tolerability of TMS. Stimulation becomes less tolerable at scalp locations near facial muscles and nerves (i.e. prefrontal locations). Low-frequency stimulation at 1 Hz is much more tolerable than 10 Hz and iTBS, while the tolerability of 10 Hz over iTBS is marginal. Researchers should consider the impact of sensation and tolerability, as well as stimulation intensity, on their study designs. Perceived sensory thresholds on the scalp can provide a rough estimate of fields strengths needed to induce neuronal effects, and qualitative descriptions may help understand the types of sensory fibers activated. Clinicians should be aware of the impact of location, frequency, and stimulation intensity on treatment tolerability. When a patient is unable to tolerate a specific parameter set, the clinician should be well-versed in possible adjustments to improve tolerability in conjunction with any data regarding the impact to efficacy. We acknowledge the limitation of a single-session study in generally healthy individuals, and there is a need for additional data to further inform research and clinical practice.

## Data Availability

All data produced in the present study may be available upon reasonable request to the authors.

## Acknowledgements

This research was not supported by any specific grant funding from agencies in the public, commercial, or non-profit sectors.

## Contributors

Neketa Nesmith: study design, data collection, data analysis, manuscript writing

Megan Senda: study design, data collection, data analysis, manuscript writing

Yang Hou: data analysis, manuscript writing

Kirthin Dev: study design, data collection

Austin M. Spitz: study design, manuscript writing

F. Andrew Kozel: study design, data analysis, manuscript writing

Kevin A. Johnson: study design, data collection, data analysis, manuscript writing

